# Sex-dependent associations between human milk oligosaccharides and malaria outcomes in breastfeeding Ugandan children

**DOI:** 10.1101/2024.03.17.24304426

**Authors:** Victor Irungu Mwangi, Tonny Jimmy Owalla, Sara Moukarzel, Emmanuel Okurut, Chloe Yonemitsu, Lars Bode, Thomas G. Egwang

**Affiliations:** Programa de Pós-Graduaçặo em Medicina Tropical, Universidade do Estado do Amazonas (UEA), Manaus, AM, Brazil; Med Biotech Laboratories, Kampala, Uganda; Department of Pediatrics, Division of Neonatology, University of California San Diego, La Jolla, California, United States of America; Larsson-Rosenquist Foundation Mother-Milk-Infant Center of Research Excellence (MOMI CORE), University of California San Diego, La Jolla, California, United States of America

**Keywords:** Human milk oligosaccharides, malaria, breastfeeding children

## Abstract

Human milk oligosaccharides (HMOs), whose compositions vary between secretor and non-secretor mothers, protect breastfeeding children against various diseases. We investigated the relationship between HMOs in Ugandan secretor mothers and malaria outcomes in their children. Malaria outcomes in breastfeeding children followed up over seven months were classified as malaria-free and asymptomatic, mild, or severe malaria. A single human milk sample collected from each mother was analyzed for HMOs. Significantly higher 2’fucosyllactose (2’FL) and lacto-N-fucopentaose I (LNFP I) concentrations were observed in mothers of malaria-free children, and significantly higher lacto-N-tetraose (LNT) concentrations were observed in mothers of children with asymptomatic malaria. Concentrations of five HMOs, 3-fucosyllactose (3 FL), 3’sialyllactose (3’SL), difucosyllactose (DFLac), lacto-N-fucopentaose II (LNFP II), and sialyllacto-*N*-tetraose b (LSTb); and two HMOs, difucosyllacto-N-tetrose (DFLNT) and fucosyllacto-N-hexaose (FLNH) were significantly higher in mothers of malaria-infected children and children with severe malaria, respectively. Sex-dependent associations were observed for some HMOs.

## Methods, Results & Discussion

Breastfeeding protects children against malaria by mechanisms which are poorly understood [1]. Human milk oligosaccharides (HMOs), representing the third most abundant component of human milk, protect breastfeeding children against diseases [2]. HMO concentrations vary between secretor and non-secretor mothers [3,4] who constitute approximately 80 % and 20 % of Ugandan mothers, respectively [5]. Secretor mothers have an active fucosyltransferase 2 (FUT2) making their milk highly abundant in FUT-2 dependent HMOs like 2’-fucosyllactose (2’FL) and lacto-N-fucopentaose I (LNFP I); non-secretor mothers with an inactive FUT2 have low milk concentrations of FUT2-dependent HMOs [1,2]. Here, we investigated the relationship between HMOs in 99 Ugandan secretor mothers and malaria outcomes in their breastfeeding children (**Supplementary Table 1**). Non-secretor mother-child pairs were not included because of the small sample size. Children were followed up between March and September 2018 for malaria infection and clinical episodes during clinic visits. Malaria diagnosis was by CareStart malaria Pf (HRP-2) rapid diagnostic test (Access Bio, NJ, USA). Malaria-free (MF) children were consistently malaria-negative during 8 clinic visits for other ailments. Children with asymptomatic malaria (AM) were malaria-positive with no symptoms or fever. Children with mild malaria (MM) were malaria-positive with non-specific symptoms like vomiting, diarrhea, poor feeding and fever (≥ 37.5 °C). Finally, children with severe malaria (SM) were malaria-positive with other symptoms including respiratory distress and severe anemia requiring blood transfusion according to the World Health Organization definition [6]. Children with AM or MM received artemether-lumefantrine (Coartem); those with SM received intravenous quinine or rectal artesunate according to national treatment guidelines.

A single human milk sample was collected from each mother in March 2018 and analyzed for 19 HMOs by high performance liquid chromatography (HPLC) as described [7]. Human milk was spiked with raffinose as an internal standard. Absolute concentrations were calculated based on standard response curves for each annotated HMO. The total concentration of HMOs was calculated as the sum of the annotated oligosaccharides and the relative abundance of each HMO was calculated. HMO-bound fucose and HMO-bound sialic acid were calculated on a molar basis. Simpson’s Diversity index was calculated based on relative abundance of HMOs. Children with laboratory-confirmed malaria were designated malaria infected (MI = AM+MM+SM; n=35); and those who experienced clinical malaria were grouped as MM+SM (n=19). Associations between HMO concentrations and malaria outcomes in all children and separately in males and females were assessed by the Mann-Whitney U and Welch t-tests.

We first compared HMOs between mothers of malaria-free (n=64) and malaria-infected (MF versus MI) children. Higher 2’FL, LNFP I, total HMOs and HMO-bound fucose (HMO-Fuc) concentrations were observed in mothers of MF children (**Figure 1A**). Although LNFP I concentrations decrease during lactation (5, 8), the median LNFP I concentration during late lactation was significantly higher in mothers of malaria-free children (1247.8 (537.4-2135.0) nmol/mL versus 450.0 (330.6-1417.2) nmol/mL; P = 0.006). By contrast, higher 3’sialyllactose (3’SL), 3-fucosyllactose (3FL), difucosyllactose (DFLac), lacto-N-fucopentaose II (LNFP II) and sialyllacto-*N*-tetraose b (LSTb) concentrations and the diversity index (reflecting variety in HMOs) were observed in mothers of MI children (**Figure 1B**). We next compared HMOs between mothers of children with AM and children with clinical malaria (AM versus MM+SM). Lacto-N-tetraose (LNT) concentrations were higher in mothers of AM children (**Figure 1C; Supplementary Table 2**). We then compared HMOs between mothers of children with mild malaria (MM) and children with severe malaria (SM). Difucosyllacto-N-tetraose (DFLNT), fucosyllacto-N-hexaose (FLNH) and HMO-Fuc concentrations were higher in mothers of SM children (**Figure 1D, Supplementary Table 3**). Finally, since MF and AM outcomes might overlap, we compared HMOs between mothers of MF children and children with AM. Diversity and LNT concentrations were higher in mothers of AM but not MF children while 2’FL, LNFPI, total HMOs and HMO-Fuc concentrations were higher in mothers of MF but not AM children (**Figure 1E and F, Supplementary Table 4**).

**Figure 1.**
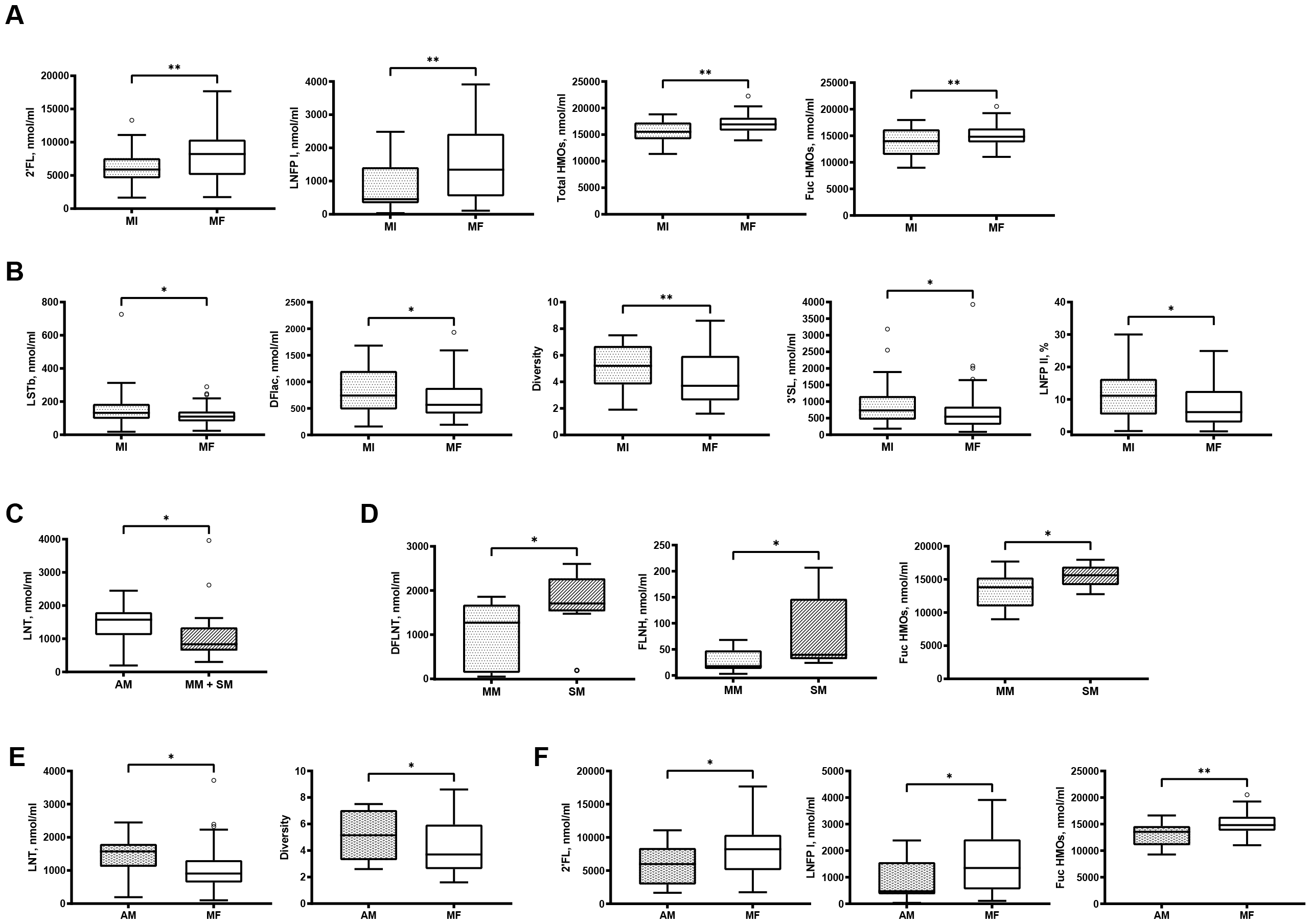
Associations between human milk oligosaccharides (HMOs) and malaria outcomes in children. **A**. Higher concentrations of α1-2 fucose-linked HMOs, total and fucose-bound HMOs in mothers of malaria-free (MF) children. **B**. Higher concentrations of α1-3- and α1-4-fucose-linked HMOs and a sialylated HMO in mothers of malaria-infected (MI) children. **C**. Higher concentrations of LNT in mothers of children with asymptomatic malaria (AM). **D**. Higher concentrations of DFLNT, FLNH and fucose-bound HMOs in mothers of children with severe malaria (SM). **E**. Higher diversity and concentrations of LNT in mothers of children with asymptomatic malaria. **F**. Higher concentrations of 2’FL, LNFP I and HMO-bound fucose in mothers of malaria-free children. Abbreviations: 2’FL, 2’fucosyllactose; LNFP I, lacto-N-fucopentaose I, 3’SL, 3’sialyllactose; DFLac, difucosyllactose; LNFPII, lacto-N-fucopentaose II (LNFP II), and LSTb, sialyllacto-*N*-tetraose b; LNT, lacto-N-tetraose; DFLNT, difucosyllacto-N-tetrose ; FLNH, fucosyllacto-N-hexaose; and Fuc HMO, HMO-bound fucose. Data are shown as box and whisker plots with the medians and the lower and upper quartiles or interquartile range (IQR). P values represent the outcomes of comparisons of HMO concentrations or diversity by malaria outcomes by either t-tests or Mann-Whitney U test. NS, non-significant; * Unadjusted P < 0.05; ** Unadjusted P < 0.01.

The relationships between HMOs and malaria outcomes were re-analyzed by children’s sex. Higher 2’FL and HMO-Fuc concentrations in mothers of MF children were observed for females (**Figure 2A**). Higher diversity and LSTb concentrations in mothers of MI children were observed for females, while higher LNFP II concentrations in mothers of MI children were observed for males (**Figure 2B**). Second, higher LNT concentrations in mothers of children with AM were observed for males while higher HMO-Fuc concentrations in mothers of children with MM+SM were also observed for males (**Figure 2C**). Finally, higher diversity and LNT concentrations in mothers of children with AM and higher LNFP I and HMO-Fuc concentrations in mothers of MF children were observed for females (**Figure 2D and E**). The analyses for all HMOs by sex are presented in **Supplementary Tables 5-7**. Analysis by sex was not done for malaria severity because of small sample sizes.

**Figure 2.**
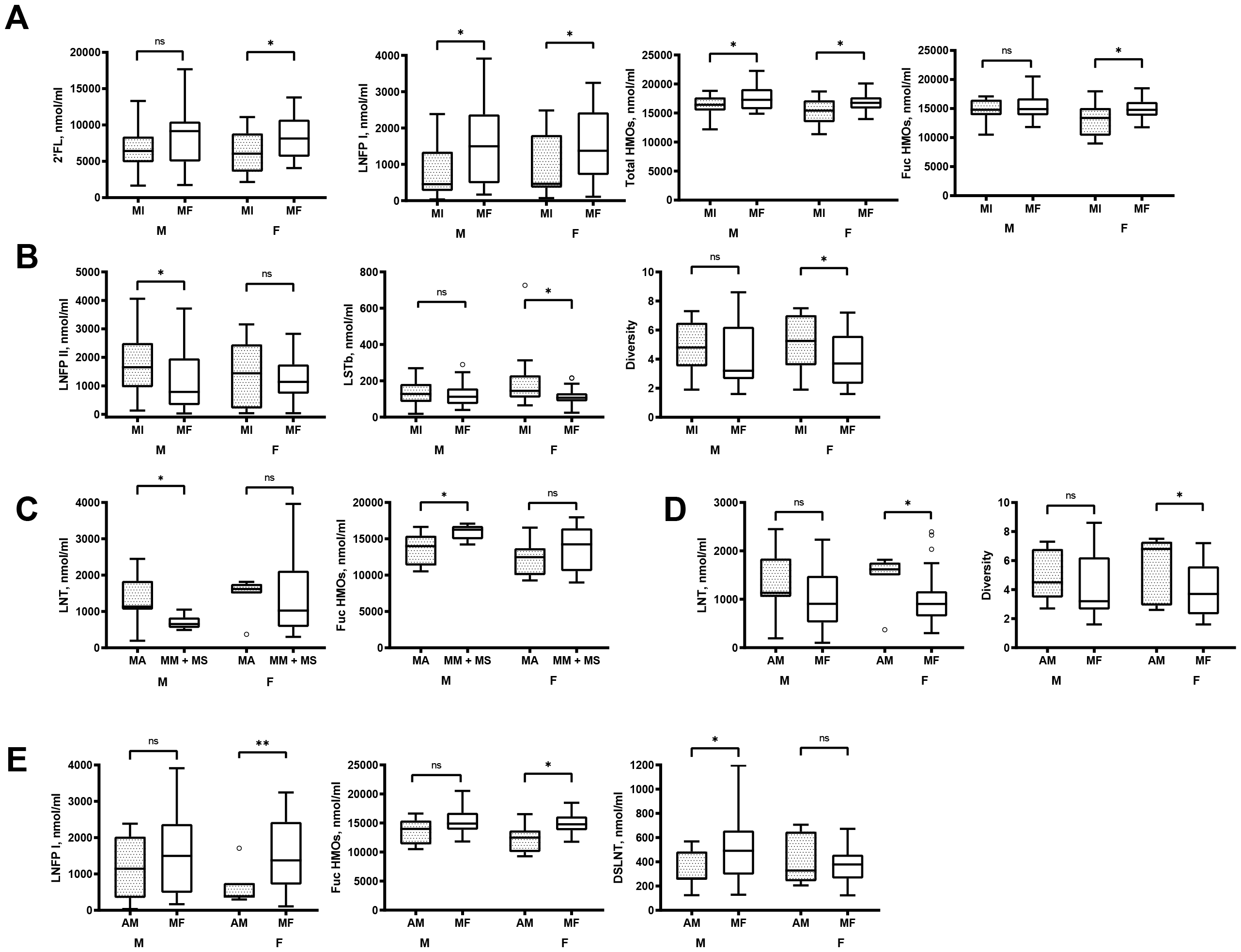
Sex-dependent associations between HMOs and malaria outcomes in children. **A**. Higher concentrations of 2’FL and fucose-bound HMOs in milk of mothers of malaria-free (MF) female children. **B**. Higher diversity and concentrations of LNFP II and LSTb in mothers of malaria-infected (MI) male and female children, respectively. **C**. Higher concentrations of LNT and fucose-bound HMOs in mothers of male children with asymptomatic (AM) and clinical malaria (MM+SM). **D**. Higher diversity and concentrations of LNT in mothers of AM female children. **E**. Higher concentrations of LNFP I, HMO-bound fucose and DSLNT in mothers of male and female children with MF status. Abbreviations: F, females; M, males. DSLNT, disialyllacto-N-tetraose; 2’FL, 2’fucosyllactose; LNFP I, lacto-N-fucopentaose I; LNFPII, lacto-N-fucopentaose II (LNFP II), and LSTb, sialyllacto-*N*-tetraose b; LNT, lacto-N-tetraose; Fuc HMO, HMO-bound fucose. Data are shown as box and whisker plots with the medians and the lower and upper quartiles or interquartile range (IQR). P values represent the outcomes of comparisons of HMO concentrations or diversity by malaria outcomes by either t-tests or Mann-Whitney U test. NS, non-significant; * Unadjusted P < 0.05; ** Unadjusted P <0.01.

It is remarkable that HMOs in milk collected at a single time point from mothers at different lactation stages were associated with malaria outcomes in children during follow up. From published [8] and our own studies [5], eight of these HMOs remain unchanged or increase in concentration throughout lactation. It is plausible that the concentrations of these HMOs at the single sampling mirrored those in mothers during follow up. Although LNFP I was reduced in late lactation [821.3 (373.8-1941.4) nmol/mL at > 6 months versus 1586.7 (620.5-2475.0) nmol/mL at ≤ 6 months; P = 0.048)], it was still associated with the malaria-free status. This raises the possibility that HMOs which decrease in early lactation may still have an effect on disease risk in breastfeeding children in late lactation.

Our data confirm previous reports that fucose-bound HMOs protected children against infectious diseases [1,2] and extends this protection to malaria. Concentrations of 2’FL and LNFP1, with an α1-2 fucose linkage, were significantly higher in mothers of malaria-free but not malaria-infected children. By contrast, LNFPII and DFLac, with α1-3 and α1-4 fucose linkages, were significantly higher in mothers of malaria-infected children. Our data support previous reports linking higher concentrations of α1-2-fucosylated HMOs to a lower incidence of diarrhea [9,10], zero sick days [11] and lower infant mortality in HIV-exposed infants [12] and higher concentrations of α1-3 fucosylated HMOs to sick days [11]. Moreover, significantly higher concentrations of LNT in mothers of children with asymptomatic malaria suggest that it might be associated with malaria clinical immunity. Additionally, high concentrations of DFLNT and FLNH were observed in mothers of children with severe but not mild malaria; previous studies have not reported associations between HMOs and disease severity in children. Finally, significant differences in HMO concentrations between mothers of children with asymptomatic malaria and malaria-free children (**Figure 1E** and **F; Figure 2D** and **E**) suggest that these malaria outcomes do not overlap in this study.

To the best of our knowledge, this is the first study to report significant sex-dependent associations between HMOs and malaria outcomes in children from a malaria-endemic region. However, the mechanisms by which HMOs influence malaria outcomes in children in a sex-dependent manner remain unknown. Some HMOs reach the systemic circulation after ingestion [13] where they engage innate immune cells implicated in malaria protection and disease [14,15,16]. HMOs also act as soluble decoy receptors which pre-empt pathogen attachment to host cells [2]. Finally, HMOs induce specific microbiota which are important for the development of immunity [2,17] and sex differences in microbiota composition [18] and innate immune responses have been reported in malaria patients [19]. Mechanistic studies of HMOs and malaria outcomes in children must be undertaken to obtain useful insights about the mechanisms of breastfeeding-mediated protection.

This study has several limitations. First, associations between HMO blood concentrations in children and malaria outcomes were not investigated. Second, the sample sizes were small especially when malaria outcomes were stratified by children’s sex and disease severity. However, this limitation does not negate the significant differences in concentrations and composition of HMOs between mothers of children with different malaria outcomes. The confirmation of prior literature linking α1-2-fucosylated HMOs and α1-3- or α1-4-fucosylated HMOs to protection and disease in children [9, 10,11, 12] validates our findings. Third, a single milk sample, rather than longitudinal milk samples, was collected from each mother at the same time point for measuring HMO concentrations. This limitation is mitigated by the observed significant associations between HMO concentrations and malaria outcomes in children and the fact that the concentrations of eight of these HMOs remain constant or increase during lactation (8). Fourth, the malaria-free status was not confirmed by a more sensitive molecular diagnostic test to distinguish it from asymptomatic malaria with low parasitemias. However, comparisons of LNT, 2’FL and LNFP I concentrations between mothers of malaria-free children and children with asymptomatic malaria suggest that these malaria outcomes do not overlap. Finally, our results are not generalizable to children of non-secretor mothers with different HMO concentrations and composition [3,4]. These limitations will be addressed in future longitudinal studies of breastfeeding secretor and non-secretor mother-child pairs.

In conclusion, our observational study demonstrated significant sex-dependent associations between malaria outcomes in breastfed children with concentrations of α1-2-fucosylated HMOs, α1-3- or α1-4-fucosylated HMOs, and sialylated HMOs in milk of Ugandan secretor mothers.

### Ethical Consideration

The Uganda National Council for Science and Technology and the Research Ethics Committee of the Vector Division, Ministry of Health, approved the original HD4MC study. All mothers who donated milk samples and brought infants to the study clinic for malaria monitoring and health assessment signed or thumb-printed an informed consent form.

## Data Availability

All data produced in the present study are available upon reasonable request to the authors

## Acknowledgements

We are grateful to the Ugandan mother-children pairs for their participation. We would like to thank the clinical and laboratory staff of Med Biotech Laboratories at St Anne HC III, Usuk subcounty in Katakwi District, Uganda.

## Financial support

Thomas Egwang received funding from the Global Innovation Fund and Grand Challenges Canada. Lars Bode is UC San Diego Chair of Collaborative Human Milk Research endowed by the Family Larsson-Rosenquist Foundation (FLRF), Switzerland. The funders played no role in the study design, data collection and analysis nor in the preparation of the manuscript and decision to publish this paper.

## Authors’ Current addresses

Victor Irungu Mwangi: Programa de Pós-Graduação em Medicina Tropical, Universidade do Estado do Amazonas (UEA), Manaus, AM, Brazil; Tonny Owalla, Emmanuel Okurut, and Thomas G. Egwang: Med Biotech Laboratories, P.O. Box 9364, Kampala, Uganda; Sara Moukarzel, Chloe Yonemitsu, and Lars Bode: University of California San Diego, La Jolla, California, United States of America. E-mail addresses.

## Notes

### Competing Interest Statement

The authors have declared no competing interest.

### Funding Statement

This study was funded by Global Innovation Fund and Grand Challenges Canada

### Author Declarations

The Uganda National Council for Science and Technology and the Research Ethics Committee of the Vector Division, Ministry of Health, approved the original HD4MC study.

## References

1. Brazeau NF, Tabala M, Kiketa L, et al. Exclusiv. Breastfeeding and Clinical Malaria Risk in 6-Month-Old Infants: A Cross-Sectional Study from Kinshasa, Democratic Republic of the Congo. Am J Trop Med Hyg. 2016;95(4):827–830

2. Bode L 2012. Huma. milk oligosaccharides: Every baby needs a sugar mama. Glycobiology 22 :1147– 62.

3. Kunz C, Meyer C, Collado MC, et al. 2017. Influenc. of Gestational Age, Secretor, and Lewis Blood Group Status on the Oligosaccharide Content of Human Milk. J Pediatr Gastroenterol Nutr. 64:789–98.

4. Totten SM, Zivkovic AM, Wu S, et al. 2012. Comprehensiv. profiles of human milk oligosaccharides yield highly sensitive and specific markers for determining secretor status in lactating mothers. J Proteome Res. 11:6124–33.

5. Owalla et al. 2023. Th. effect of non-genetic determinants of human milk oligosaccharide profiles in milk of Ugandan mothers. MS submitted.

6. Imbert P, Gérardin P, Rogier C, et al. 2002. Sever. falciparum malaria in children: a comparative study of 1990 and 2000 WHO criteria for clinical presentation, prognosis and intensive care in Dakar, Senegal. Trans R Soc Trop Med Hyg. 96:278–81.

7. Berger PK, Hampson HE, Schmidt KA, et al. 2023. Stabilit. of Human-Milk Oligosaccharide Concentrations Over 1 Week of Lactation and Over 6 Hours Following a Standard Meal. J Nutr. 14;152:2727–33.

8. Plows JF, Berger PK, Jones RB, et al. 2021. Longitudina. Changes in Human Milk Oligosaccharides (HMOs) Over the Course of 24 Months of Lactation. J Nutr. 151:876–82.

9. Newburg DS, Ruiz-Palacios GM, Altaye M, et al. 2004. Innat. protection conferred by fucosylated oligosaccharides of human milk against diarrhea in breastfed infants. Glycobiology. 14:253–63.

10. Morrow AL, Ruiz-Palacios GM, Altaye M, et al. 2004. Huma. milk oligosaccharide blood group epitopes and innate immune protection against campylobacter and calicivirus diarrhea in breastfed infants. Adv Exp Med Biol. 554:443–46.

11. Davis JCC, Lewis ZT, Krishnan S, et al. 2017. Growt. and Morbidity of Gambian Infants are Influenced by Maternal Milk Oligosaccharides and Infant Gut Microbiota. Sci Rep. 7:40466.

12. Kuhn L, Kim H-Y, Hsiao L, et al. 2015. Oligosaccharid. composition of breast milk influences survival of uninfected children born to HIV-infected mothers in Lusaka, Zambia. J Nutr. 145:66–72.

13. Goehring KC, Kennedy AD, Prieto PA, Buck RH. 2014. Direc. evidence for the presence of human milk oligosaccharides in the circulation of breastfed infants. PLoS One. 2014;9(7):e101692.

14. Triantis V, Bode L and van Neerven RJJ. 2018. Immunologica. Effects of Human Milk Oligosaccharides. Front. Pediatr. 6:190.

15. Chua CL, Brown G, Hamilton JA, Rogerson S, Boeuf P. 2013. Monocyte. and macrophages in malaria: protection or pathology? Trends Parasitol. 29:26–34.

16. Dobbs KR, Crabtree JN, Dent AE. 2020. Innat. immunity to malaria-The role of monocytes. Immunol Rev. 293:8–24.

17. Berger B, Porta N, Foata F, et al. 2020. Linkin. Human Milk Oligosaccharides, Infant Fecal Community Types, and Later Risk To Require Antibiotics. mBio. 2020;11(2):e03196–19.

18. Valeri F, Endres K. 2021. Ho. biological sex of the host shapes its gut microbiota. Front Neuroendocrinol. 61:100912.

19. Kanoi BN, Egwang TG. 2021. Se. differences in concentrations of HMGB1 and numbers of pigmented monocytes in infants and young children with malaria. Parasitol Int. 84:102387.

