## Supplementary Table 1 for "Sex-dependent associations between human milk oligosaccharides and malaria outcomes in breastfeeding Ugandan children"

**Supplementary Table 1. Characteristics of secretor mother-child study population**

| **Characteristic** | Mean (SD) | Median (IQR) | MF | AM | MM | SM |
|---|---|---|---|---|---|---|
| **Children** | | | n=56 | n=16 | n=9 | n=7 |
| Age (Weeks) | 45.9 (24.2) | 46.0 (28.0 - 66.0) | 41.9 (23.2) | 48.4 (20.6) | 61.9 (30.6) | 59.3 (20.2) |
| Birthweight (Kg) | 3.1 (0.6) | 3.1 (2.9 - 3.5) | 3.1 (0.7) | 3.2 (0.5) | 3.1 (0.5) | 3.3 (0.5) |
| Weight (Kg) | 8.6 (1.7) | 9.0 (7.0 - 10.0) | 8.3 (1.6) | 9.0 (1.6) | 9.2 (1.8) | 10.1 (1.3) |
| Hemoglobin (g/dL) | 10.9 (1.2) | 11.0 (10.3 - 11.8) | 10.9 (1.0) | 10.6 (1.2) | 11.1 (2.0) | 11.4 (0.9) |
| Sex | | | | | | |
| Male | | | 24 | 9 | 3 | 4 |
| Female | | | 31 | 7 | 6 | 3 |
| Clinic visits (range) | | | 8* | 8 | 8 | 8 |
| Malaria episodes (#) | | | 0 | 22 | 19 | 10 |
| **Mothers** | | | N=63 | N=16 | N=10 | N=9 |
| Age (years) | 26.4 (6.6) | 25.0 (21.0 - 30.0) | 26.0 (6.1) | 25.3 (6.5) | 28.3 (7.3) | 29.0 (8.7) |
| BMI (kg/m$^2$) | 23.3 (2.1) | 23.2 (22.0 - 24.4) | 23.3 (2.2) | 23.0 (1.8) | 23.7 (2.4) | 23.1 (1.5) |
| MUAC (cm) | 26.8 (2.0) | 26.8 (25.4 - 28.0) | 26.9 (2.1) | 26.5 (2.1) | 27.6 (1.6) | 26.2 (1.7) |
| Lactation duration (weeks) | 40.8 (23.9) | 45.0 (23.0 - 60.0) | 37.2 (23.3) | 43.3 (21.0) | 55.7 (23.7) | 44.8 (24.8) |

**Legend**: * Clinic visits for other ailments and consistently tested malaria-negative; SD - Standard deviation; IQR - interquartile range; BMI - Body mass index; MUAC - mid upper arm circumference.

**Supplementary Table 2: Median (IQR) or mean (SD) concentrations of HMO concentrations in mothers of children with asymptomatic malaria and clinical malaria**

| HMO (%) | AM (n=16) Median (IQR) | MM + SM (n=19) Median (IQR) | p-value† |
|---|---|---|---|
| 2'FL | 36.45 (22.15 - 51.48) | 37.10 (31.40 - 43.90) | 0.6294 |
| 3'FL | 2.65 (2.00 - 4.23) | 3.40 (2.20 - 5.00) | 0.3204 |
| LNnT * | 6.06 (2.57) | 5.64 (2.40) | 0.6229 |
| 3'SL | 4.00 ( 3.23 - 5.03) | 5.20 ( 2.70 - 8.20) | 0.1794 |
| DFlac | 3.90 ( 3.23 - 9.40) | 5.90 ( 3.80 - 7.70) | 0.2395 |
| 6'SL | 0.80 (0.63 - 1.75) | 0.90 (0.60 - 1.50) | 0.9804 |
| **LNT** | 10.85 (6.95 - 12.95) | 5.00 (3.80 - 8.60) | **0.0432** |
| LNFP I | 3.55 (2.63 - 9.43) | 2.90 (1.90 - 7.60) | 0.7622 |
| LNFP II * | 12.34 (9.25) | 9.832 (6.30) | 0.3656 |
| LNFP III | 0.45 (0.30 - 0.85) | 0.40 (0.30 - 0.50) | 0.4688 |
| LSTb | 1.00 (0.53 - 1.25) | 0.90 (0.70 - 1.20) | 0.9804 |
| LSTc | 0.30 (0.13 - 0.50) | 0.30 (0.10 - 0.50) | 0.8748 |
| DFLNT | 8.35 (1.63 - 12.70) | 9.60 (1.10 - 11.90) | 0.4362 |
| LNH | 0.55 (0.35 - 0.85) | 0.40 (0.20 - 0.60) | 0.1652 |
| DSLNT * | 2.54 (1.23) | 2.74 (1.09) | 0.6205 |
| FLNH | 0.25 (0.10 - 0.40) | 0.20 (0.10 - 0.40) | 0.9036 |
| DFLNH | 0.20 (0.10 - 0.28) | 0.10 (0.10 - 0.40) | 0.8054 |
| FDSLNH | 1.20 ( 0.48 - 2.20) | 0.20 (0.10 - 1.80) | 0.2405 |
| DSLNH | 0.15 (0.10 - 0.35) | 0.10 (0.10 - 0.30) | 0.9791 |

**Legend**: AM, children with asymptomatic malaria; MM+SM, children with clinical malaria; %, percentage. The asterisk (*) denotes HMOs with a parametric distribution analyzed by unpaired t-test with the Welch correction. Dagger sign (†) indicates that the p-values are from unadjusted analysis. For the parametric distributions, we used the mean (SD) as presented in the table. HMO concentrations here are in %.

**Supplementary Table 3: Median (IQR) or mean (SD) HMO concentrations in mothers of children with mild malaria (MM) and mothers of children with severe malaria (SM)**

| HMO (%) | MM (n=10) Median (IQR) | SM (n=9) Median (IQR) | p-value† |
|---|---|---|---|
| 2'FL * | 38.53 (14.42) | 41.70 (12.98) | 0.6206 |
| 3'FL | 3.30 (1.28 - 5.15) | 3.40 (3.00 - 4.85) | 0.7650 |
| LNnT * | 6.19 (3.13) | 5.03 (1.08) | 0.2950 |
| 3'SL | 4.80 (1.58 - 8.03) | 6.40 (4.90 - 9.00) | 0.1895 |
| DFlac * | 5.71 (2.64) | 5.79 (2.00) | 0.9420 |
| 6'SL | 0.85 (0.48 - 1.15) | 1.10 (0.85 - 1.80) | 0.2331 |
| LNT * | 8.62 (6.82) | 5.44 (2.04) | 0.1887 |
| LNFP I | 3.55 ( 1.35 - 13.63) | 2.90 (1.90 - 7.25) | 0.7627 |
| LNFP II * | 10.46 (7.64) | 9.13 (4.75) | 0.6525 |
| LNFP III * | 0.45 (0.17) | 0.40 (0.10) | 0.4319 |
| LSTb * | 1.02 (0.41) | 0.83 (0.39) | 0.3261 |
| LSTc | 0.25 ( 0.175 - 0.35) | 0.30 (0.10 - 0.70) | 0.6995 |
| DFLNT | 8.85 (0.93 - 10.35) | 11.90 (9.35 - 13.35) | 0.0503 |
| LNH | 0.40 (0.20 - 0.65) | 0.40 (0.30 - 0.90) | 0.7688 |
| DSLNT * | 3.17 (1.27) | 2.27 (0.62) | 0.0666 |
| **FLNH** | 0.10 (0.08 - 0.33) | 0.30 (0.20 - 0.95) | **0.0314** |
| DFLNH | 0.1000 (0.10 - 0.53) | 0.20 (0.10 - 0.40) | 0.6213 |
| FDSLNH | 0.20 (0.10 - 1.95) | 0.90 (0.10 - 2.00) | 0.8522 |
| DSLNH * | 0.16 (0.14) | 0.31 (0.31) | 0.2092 |

**Legend**: MM, children with mild malaria; SM, children with severe malaria; %, percentage. The asterisk (*) denotes HMOs with a parametric distribution analyzed by unpaired t-test with the Welch correction. Dagger sign (†) indicates that the p-values are from unadjusted analysis. For the parametric distributions, we used the mean (SD) as presented in the table. HMO concentrations here are in %.

**Supplementary Table 4:** Median (IQR) or mean (SD) concentrations of HMOs in mothers of children with asymptomatic malaria or malaria-free status

| HMO (nmol/mL) | MF (n=64) | AM (n=16) | p value† |
|---|---|---|---|
| **2’FL** * | 8238 (3285) | 5868 (2995) | **0.0103** |
| 3'FL | 337.9 (284.7 - 594.7) | 401.1 (313.9 - 562.8 | 0.3230 |
| LNnT | 768.9 (515.4 - 1100) | 805.4 (700.9 - 1015) | 0.4992 |
| 3’SL | 544.9 (300.4 - 848.5) | 604.7 (406.7 - 785.4) | 0.4055 |
| DFlac | 567.8 (403.2 - 886.5) | 613.0 (468.6 - 1003) | 0.4188 |
| 6’SL | 149.5 (99.0 - 289.8) | 131.9 (100.8 - 270.9) | 0.7689 |
| **LNT** | 909.6 (639.3 - 1313) | 1575 (1111 - 1802) | **0.0148** |
| **LNFP I** | 1347 (542.9 - 2426) | 474.7 (359.2 - 1567) | **0.0262** |
| LNFP II | 1074 (541.4 - 1962) | 1519 (853.6 - 2774) | 0.0981 |
| LNFP III | 67.3 (49.9 - 82.4) | 70.0 (47.1 - 107.4) | 0.5857 |
| LSTb | 109.6 (81.1 - 140.5) | 130.3 (85.9 - 183.6) | 0.1477 |
| LSTc | 54.3 (24.7 - 129.6) | 50.1 (19.9 - 73.1) | 0.4362 |
| DFLNT | 896.2 (78.0 - 1579) | 1128 (285.3 - 1759) | 0.5458 |
| LNH | 61.6 (33.8 - 99.0) | 82.1 (57.3 - 128.3) | 0.1412 |
| DSLNT | 412.1 (261.2 - 506.6) | 319.2 (251.3 - 523.4) | 0.4911 |
| FLNH | 24.5 (9.1 - 50.5) | 35.3 (12.7 - 62.5) | 0.3531 |
| DFLNH | 39.0 (13.9 -77.4) | 25.6 (12.1 - 44.8) | 0.3141 |
| FDSLNH | 146.9 (30.9 - 261.9) | 182.4 (74.3 - 318.1) | 0.3919 |
| DSLNH | 40.5 (15.2 - 64.0) | 21.0 (12.8 - 56.2) | 0.2573 |
| **Total HMO** * | 17012 (1739) | 15284 (2139) | **0.0071** |
| Sia HMO | 2275 (1869 - 2765) | 2318 (1950 - 2544) | 0.9842 |
| **Fuc HMO** * | 15107 (2019) | 13119 (2329) | **0.0050** |
| **Diversity** | 3.7 (2.6 - 6.0) | 5.2 (3.3 - 7.1) | **0.0232** |

**Legend:** AM, asymptomatic malaria; MF, malaria-free; nmol/mL, nanomole per milliliter. The asterisk (*) denotes HMOs with a parametric distribution analyzed by unpaired t-test with the Welch correction. Dagger sign (†) indicates that the p-values are from unadjusted analysis. For the parametric distributions, we used the mean (SD) as presented in the table.

**Supplementary Table 5: Median (IQR) or mean (SD) concentrations of HMOs in mothers of male and female children who are malaria-infected or malaria-free**

| | MALE | | | FEMALE | | |
|---|---|---|---|---|---|---|
| **HMO (nmol/mL)** | **MI (n=16)** | **MF (n=24)** | **p-value†** | **MI (n=16)** | **MF (n=31)** | **p-value†** |
| **2’FL** * | 6671 (2935) | 8738 (3860) | 0.0626 | 6084 (2856 | 8223 (2941) | **0.0221** |
| 3'FL | 487.6 (357.4 - 817.8) | 337.8 (294.5 - 629.5) | 0.0783 | 403.7 (267.7 - 729.2) | 327.1(260.0 - 475.8) | 0.3792 |
| LNnT | 762.3 (597.4 - 984.6) | 879.0 (487.0 - 1141) | 0.7693 | 847.2 (746.7 - 1082) | 762.0 (439.5 - 1084) | 0.1691 |
| 3’SL | 737.8 (468.2 - 1455) | 544.9 (281.5 - 882.4) | 0.0560 | 604.7 (312.6 - 980.6) | 403.4 (300.4 - 801.7) | 0.3106 |
| DFlac * | 946.4 (928.5) | 567.8 (689.2) | 0.0685 | 735.2 ( 479.0 - 1148) | 462.7 (378.9 - 745.6) | 0.0957 |
| 6’SL | 155.1 (91.3 - 270.9) | 201.1 (100.3 - 511.9) | 0.2523 | 141.0 (96.3 - 198.2) | 123.5 (82.3 - 180.5) | 0.7140 |
| LNT | 1084 (607.6) | 1036 (586.9) | 0.8076 | 1513 (724.2 - 1726) | 903.0 (646.1 - 1167) | 0.0993 |
| **LNFP I** | 455.9 (266.8 - 1349) | 1499 (480.4 - 2376) | **0.0437** | 466.2 (359.2 - 1807) | 1375 (709.9 - 2430) | **0.0156** |
| **LNFP II*** | 1652 (953.0 - 2504) | 787.8 (321.9 - 1962) | **0.0468** | 1409 (1082) | 1199 (826.8) | 0.5026 |
| LNFP III | 61.7 (47.10 - 84.2) | 74.5 (54.1 - 92.0) | 0.3011 | 62.60 (54.2 - 77.8) | 60.30 (44.5 - 78.9) | 0.3883 |
| **LSTb** | 130.5 (64.9) | 122.5 (62.4) | 0.7010 | 145.1 (107.4 - 230.6) | 107.0 (86.7 - 131.5) | **0.0187** |
| LSTc | 44.2 (22.4 - 73.1) | 73.8 (30.7 - 200.3) | 0.1588 | 40.8 (19.9 - 59.5) | 41.8 (18.3 - 65.5) | 0.7682 |
| DFLNT | 1577 (351.0 - 1937) | 964.1 (67.8 - 1484) | 0.0737 | 1255 (142.7 - 1739) | 496.3 (58.5 - 1838) | 0.5410 |
| LNH | 71.6 (43.9 - 82.5) | 54.4 (22.6 - 109.1) | 0.3557 | 82.3 (44.3 - 127.2) | 61.1 (37.2 - 90.8) | 0.4427 |
| DSLNT * | 383.0 (137.5) | 506.3 (253.0) | 0.0543 | 436.0 (219.2) | 378.6 (120.5) | 0.3415 |
| FLNH | 26.6 (14.4 - 58.5) | 21.4 (9.3 - 58.2) | 0.6973 | 40.8 (15.5 - 65.4) | 21.5 (7.5 - 49.7) | 0.2008 |
| DFLNH | 15.4 (8.3 - 56.3) | 41.0 (18.1 -76.2) | 0.0938 | 26.7 (15.1 - 74.1) | 41.1 (16.9 - 78.5) | 0.6852 |
| FDSLNH | 182.4 (37.7 - 290.2) | 95.9 (24.7 - 206.2) | 0.3277 | 25.5 (13.1 - 294.5) | 168.4 (31.6 - 274.0) | 0.5229 |
| DSLNH | 25.4 (12.8 - 56.2) | 40.6 (11.5 - 92.2) | 0.3485 | 18.8 (9.2 - 41.2) | 24.5 (6.4 - 60.0) | 0.3299 |
| **Total HMO** * | 16243 (1845) | 17615 (1998) | **0.0325** | 15355 (2189) | 16653 (1479) | **0.0442** |
| Sia HMO | 2591 (908.4) | 2658 (843.3) | 0.8146 | 2375 (1955 - 2786) | 2208 (1579 - 2492) | 0.2153 |
| **Fuc HMO** * | 14666 (2086) | 15497 (2326) | 0.2467 | 13136 (2875) | 14897 (1656) | **0.0347** |
| **Diversity** | 4.8 (3.5 - 6.5) | 3.2 (2.6 - 6.2) | 0.1626 | 5.3 (3.6 - 7.0) | 3.7 (2.3 - 5.6) | **0.0342** |

**Legend:** MI, malaria-infected children; MF, malaria-free children; nmol/mL, nanomole per milliliter. The asterisk (*) denotes HMOs with a parametric distribution analyzed by unpaired t-test with the Welch correction. Dagger sign (†) indicates that the p-values are from unadjusted analysis. For the parametric distributions, we used the mean (SD) as presented in the table.

**Supplementary Table 6: Median (IQR) or mean (SD) concentrations of HMOs in mothers of male and female children with asymptomatic or clinical malaria**

| | MALE | | | FEMALE | | |
|---|---|---|---|---|---|---|
| **HMO (nmol/mL)** | **AM (n= 9)** | **MM + SM (n= 7)** | **p-value†** | **AM (n= 7)** | **MM + SM (n= 9)** | **p-value†** |
| 2'FL * | 6160 (2960) | 7328 (2992) | 0.4507 | 5492 (3232) | 6545 (2629) | 0.4978 |
| 3'FL * | 400.8 (323.6 - 610.8 | 788.8 (476.5 - 908.7) | 0.1075 | 439.0 (181.3) | 562.4 (445.4) | 0.4664 |
| LNnT * | 968.0 (528.8) | 699.1 (250.8) | 0.2040 | 854.0 (232.0) | 1056 (503.1) | 0.3072 |
| 3'SL * | 630.0 (412.8 - 794.2) | 1197 (734.0 - 1744) | 0.0668 | 676.0 (404.9) | 873.0 (751.3) | 0.5140 |
| DFlac * | 795.6 (431.2) | 1099 (348.4) | 0.1415 | 700.9 (304.8) | 866.8 (417.5) | 0.3740 |
| 6'SL * | 206.0 (126.3) | 152.4 (98.29) | 0.3557 | 102.2 (94.90 - 151.6) | 144.3 (99.05 - 242.3) | 0.6065 |
| **LNT *** | 1384 (658.2) | 697.2 (188.9) | **0.0144** | 1618 (1494 - 1759) | 1024 (574.3 - 2122) | 0.4698 |
| LNFP I | 1145 (340.7 - 2028) | 333.7 (245.5 - 599.3) | 0.2863 | 378.3 ( 358.1 - 750.0) | 1231 (338.8 - 2043) | 0.2991 |
| LNFP II* | 1848 (1293) | 1642 (922.6) | 0.7149 | 1636 (1138) | 1233 (1069) | 0.4835 |
| LNFP III | 60.50 (46.5 - 117.4) | 62.80 (58.90 - 71.1) | 0.9769 | 77.20 (53.90 - 108.5) | 60.50 (52.60 - 69.1) | 0.2991 |
| LSTb * | 133.9 (67.7) | 126.2 (66.1) | 0.8227 | 142.8 (105.7 - 313.1) | 147.3 (103.6 - 229.9) | 0.9182 |
| LSTc | 60.50 (31.50 - 82.8) | 40.30 (15.20 - 46.7) | 0.1651 | 29.80 (18.50 - 58.3) | 42.60 (21.35 - 68.7) | 0.6065 |
| DFLNT * | 1041 ( 813.2) | 1611 (721.6) | 0.1652 | 1167 (605.1) | 1073 (967.6) | 0.9182 |
| LNH | 81.4 (54.7 - 95.2) | 61.5 (38.5 - 71.9) | 0.2004 | 103.9 (60.8) | 74.8 (55.9) | 0.3425 |
| DSLNT * | 343.8 (147.9) | 433.4 (113.2) | 0.1913 | 423.5 (201.9) | 445.7 (243.4) | 0.8448 |
| FLNH | 15.8 (9.9 - 58.5) | 29.0 (16.9 - 67.9) | 0.2861 | 58.2 (27.9 - 74.1) | 39.6 (10.3 - 55.0) | 0.4079 |
| DFLNH | 25.9 (9.9 - 66.2) | 11.6 (7.9 - 66.1) | 0.5179 | 25.3 (12.1 - 50.4) | 32.7 (18.2 - 76.8) | 0.4698 |
| FDSLNH * | 183.7 (147.2) | 177.3 (149.0) | 0.9337 | 239.0 (12.6 - 420.2) | 17.0 (11.8 - 138.8) | 0.1738 |
| DSLNH * | 30.3 (21.2) | 33.5 (29.3) | 0.8092 | 18.0 (11.6 - 84.7) | 19.2 (7.5 - 38.3) | 0.9999 |
| Total HMO * | 15885 (2052) | 16702 (1567) | 0.3819 | 14512 (2143) | 16011 (2105) | 0.1853 |
| Sia HMO | 1996 (1873 - 2463) | 2796 (2335 - 3501) | 0.1341 | 2348 (2209 - 2644) | 2402 (1785 - 3002) | 0.8371 |
| **Fuc HMO** * | 13679 (2195) | 15936 (1054) | **0.0190** | 12400 (2461) | 13708 (3181) | 0.3692 |
| Diversity* | 4.9 (1.7) | 4.7 (1.6) | 0.7452 | 5.6 (2.1) | 4.8 (1.6) | 0.4105 |

**Legend:** AM, asymptomatic malaria; MM, mild malaria; SM, severe malaria; nmol/mL, nanomole per milliliter. The asterisk (*) denotes HMOs with a parametric distribution analyzed by unpaired t-test with the Welch correction. Dagger sign (†) indicates that the p-values are from unadjusted analysis. For the parametric distributions, we used the mean (SD) as presented in the table.

**Supplementary Table 7: Median (IQR) or mean (SD) concentrations of HMOs in mothers of female and male children with asymptomatic malaria or malaria-free status.**

| | MALE | | | FEMALE | | |
|---|---|---|---|---|---|---|
| **HMO (nmol/mL)** | **AM (n=9)** | **MF (n=24)** | **p-value†** | **AM (n=7)** | **MF (n=31)** | **p-value†** |
| 2'FL * | 6160 (2960) | 8738 (3860) | 0.0555 | 5492 (3232) | 8223 (2941) | 0.0726 |
| 3'FL | 400.8 (323.6 - 610.8) | 337.8 (294.5 - 629.5) | 0.4326 | 401.3 (271.1 - 584.1) | 327.1(260.0 - 475.8) | 0.4599 |
| LNnT * | 968.0 (528.8) | 868.0 (386.1) | 0.6146 | 854.0 (232.0) | 790.2 (375.3) | 0.5732 |
| 3'SL | 630.0 (412.8 - 794.2) | 544.9 (281.5 - 882.4) | 0.5862 | 584.5 (364.9 - 801.3) | 403.4 (300.4 - 801.7) | 0.5303 |
| DFlac * | 795.6 (431.2) | 689.2 (354.1) | 0.5208 | 631.2 (457.1 - 830.3) | 462.7 (378.9 - 745.6) | 0.3382 |
| 6'SL | 155.3 (107.1 - 273.7) | 201.1 (100.3 - 511.9) | 0.5588 | 102.2 (94.9 - 151.6) | 123.5 (82.3 - 180.5) | 0.9121 |
| LNT | 1384 (658.2) | 1036 (586.9) | 0.1871 | 1618 (1494 - 1759) | 903.0 (646.1 - 1167) | **0.0295** |
| LNFP I * | 1101 (882.1) | 1552 (1102) | 0.3443 | 378.3 (358.1 - 750.0) | 1375 (709.9 - 2430) | **0.0082** |
| LNFP II* | 1106 (968.0 - 3007) | 787.8 (321.9 - 1962) | 0.0703 | 1636 (1138) | 1199 (826.8) | 0.3670 |
| LNFP III | 60.5 (46.5 - 117.4) | 74.5 (54.1 - 92.0) | 0.6418 | 77.2 (53.9 - 108.5) | 60.3 (44.5 - 78.9) | 0.2053 |
| LSTb * | 133.9 (67.7) | 122.5 (62.4) | 0.6682 | 142.8 (105.7 - 313.1) | 107.0 (86.7 - 131.5) | 0.1053 |
| LSTc | 60.5 (31.5 - 82.8) | 73.8 (30.7 - 200.3) | 0.4326 | 29.8 (18.5 - 58.3) | 41.8 (18.3 - 65.5) | 0.5841 |
| DFLNT | 1094 (86.7 - 1852) | 964.1 (67.8 - 1484) | 0.4810 | 1161 (978.2 - 1766) | 496.3 (58.5 - 1838) | 0.6057 |
| LNH | 81.4 (54.7 - 95.2) | 54.4 (22.6 - 109.1) | 0.2510 | 93.0 (52.6 - 141.6) | 61.1 (37.2 - 90.8) | 0.2193 |
| DSLNT * | 343.8 (147.9) | 506.3 (253.0) | **0.0317** | 423.5 (201.9) | 378.6 (120.5) | 0.5889 |
| FLNH | 15.8 (9.9 - 58.5) | 21.4 (9.3 - 58.2) | 0.9289 | 58.2 (27.9- 74.1) | 21.5 (7.5 - 49.7) | 0.0905 |
| DFLNH | 25.9 (9.9 - 66.2) | 41.0 (18.1 -76.2) | 0.2860 | 25.3 (12.1 - 50.4) | 41.1 (16.9 - 78.5) | 0.4341 |
| FDSLNH | 171.4 (83.6 - 252.0) | 95.9 (24.7 - 206.2) | 0.3870 | 239.0 (12.6 - 420.2) | 168.4 (31.6 - 274.0) | 0.3514 |
| DSLNH | 30.7 (12.9 - 48.7) | 40.6 (11.5 - 92.2) | 0.4565 | 18.0 (11.6 - 84.7) | 24.5 (6.4 - 60.0) | 0.6016 |
| Total HMO * | 15885 (2052) | 17615 (1998) | **0.0474** | 14512 (2143) | 16653 (1479) | **0.0388** |
| Sia HMO | 1996 (1873 - 2463) | 2403 (2038 - 3305) | 0.4454 | 2348 (2209 - 2644) | 2208 (1579 - 2492) | 0.2339 |
| Fuc HMO * | 13679 (2195) | 15497 (2326) | 0.0544 | 12400 (2461) | 14897 (1656) | **0.0365** |
| Diversity | 4.5 (3.5 - 6.8) | 3.2 (2.6 - 6.2) | 0.1564 | 6.8 (2.9 -7.3) | 3.7 (2.3 - 5.6) | **0.0353** |

**Legend:** AM, asymptomatic malaria; MF, malaria-free; nmol/mL, nanomole per milliliter. The asterisk (*) denotes HMOs with a parametric distribution analyzed by unpaired t-test with the Welch correction. Dagger sign (†) indicates that the p-values are from unadjusted analysis. For the parametric distributions, we used the mean (SD) as presented in the table.
